# Early prediction and fairness evaluation of perinatal depression using EHR: A study of 18,000+ Pregnancies

**DOI:** 10.1101/2025.06.19.25329946

**Authors:** Varuni Sarwal, Aditya Pimplaskar, Misty Richards, Kunmi Sobowale, Jeffrey N. Chiang, Loes Olde Loohuis

## Abstract

Perinatal depression (PND), defined as a depressive illness occurring during pregnancy or following childbirth, affects between 10-20% of mothers. It is one of the greatest causes of mortality and morbidity in mothers and is associated with poor outcomes in children. Early identification of at-risk mothers has the potential to greatly reduce its impact. While specific risk factors for PND have been identified, most notably a history of prior depression, it is unclear whether mothers’ Electronic Health Records (EHR) can be used early in pregnancy to predict who will go on to develop PND, especially in mothers without a history of prior depression. In this paper, we used clinical EHR data from the UCLA health system to develop predictive models of perinatal depression at a patients’ first prenatal visit (n = 18,081 pregnant mothers, n=4,307 with PND). We used a variety of predictive models, including Ridge Regression, Gradient Boosting Trees, Random Forests, and ExtraTrees. We performed separate analyses including only mothers without a history of prior depression. We further evaluated the robustness and fairness of our algorithms comparing models stratified by self-reported ethnoracial group and social determinants of health (e.g., social vulnerability index (SVI)). All model architectures used perform similarly. The Random Forest model provided robust performance with the highest accuracy and well-balanced sensitivity and specificity (AUROC 0.75, CI [0.66,0.84] in the full cohort). However, performance was reduced among mothers without prior depression (AUROC 0.71, CI [0.6,0.8]). Important risk factors identified by our model include known risk factors, such as prior mental health histories (prior depression, anxiety disorders), socioeconomic factors (social vulnerability), patient vitals (blood pressure), and measures of inflammation in blood (white blood cell counts, platelet counts), as well as novel ones (patient pulse, mean platelet volume (MPV), red blood cell distribution width (RDWSD) and rapid plasma reagin (RPR)). We observed similar model performance when stratifying our cohort by social determinants of health, with overlapping ROC bounds, equalized odds ratios between groups close to 0.8, and largely overlapping predictors of importance across models. This was not the case for ethnoracial groups, where despite observing top predictive features varied by ethnoracial category.

## Introduction

Perinatal depression (PND) is defined as depression occurring either during pregnancy or up to a year following childbirth. It is one of the greatest causes of mortality and morbidity in mothers, including a high risk of suicide and long term mental health consequences^1–3^. PND has also been associated with poor outcomes for the child, such as greater use of the pediatric emergency department, language, cognitive, emotional, and social developmental delays^2–6^. PND occurs in women of reproductive age with onset coupled with pregnancy and childbirth, a time in which the body undergoes tremendous change. While several individual biological and psychosocial risk factors have been identified, including previous psychiatric disorders like depression, anxiety^7^ and ADHD^8^, complications at birth^9^, inflammatory markers, social factors such as family and partner support^10^, adverse life events^11^, and environmental factors such as job status, *integrated predictive models* that have the potential to yield the greatest clinical impact are urgently called for^12–14^. Several recent studies have focused on building such models, using non-clinical data such as sleep^15–23^ self-reported smartphone app data^15–24^, social and behavioral data such as worry and maternal adverse childhood experiences^25^ or have focused on specific high stress global pandemic situations (COVID-19^15–23^).

Recently, prediction of PND from electronic health records (EHR) and registry data has recently started receiving increased attention. The majority of studies published so far focus on the problem of postpartum depression, using information gathered throughout pregnancy^18,26–35^. This lack of inclusion of depression *during pregnancy* is surprising, given that the onset of PND is most frequently *prior to* giving birth^36,37^, and, if left untreated, naturally and frequently continues into the postpartum period^36–39^.

In this work, we aim to predict PND, broadly defined as an onset of depression or antidepressant medication initiation during pregnancy or up to one year after delivery, using EHR data available at/around the first prenatal visit. To do so, we use the UCLA health records from >18,000 pregnant mothers, and build a prediction model based on information available at the first prenatal visit, prior to depressive onset. UCLA serves a diverse patient population, and rich records are available, which include social determinants of health: i.e., social vulnerability index (SVI)^40^, Healthy Places Index (HPI)^41^, and Barriers to Accessing Service (BAS) score^42^, for all patients. We evaluate a variety of machine learning models both including and excluding mothers with a prior history of depression or mental health-related diagnoses. Further, our diverse patient population enables us to evaluate the *fairness* of predicting PND across ethnoracial categories, and across social determinants of health strata.

## Methods

### Dataset

We extracted EHR from UCLA Health of patients from January 2011 to September 2024 that were de-identified for research purposes. This dataset contained births associated with prenatal visits. EHR data was extracted for all patients with ICD codes denoting pregnancy (Z37.0/1/2/3/4/5/6/7/8).

### Cohort definition and case assignment

In our cohort, all pregnant mothers whose pregnancy was associated with a live birth, and who had an initial prenatal or routine prenatal visit before gestational age of 30 weeks were included. For each woman with multiple births, we included only the pregnancy associated with their first birth. PND cases were defined as women with a pregnancy associated with a live birth and (1) diagnosis of depression, postpartum depression or related codes, namely F53.0, F32.A, F32.89, F32.9, O99.344, O99.345, O99.343, O99.340 and others (Figure 1); or (2) new treatment with antidepressant drug (complete list in Table S2), during pregnancy or within 1 year of childbirth. We further excluded mothers who had received diagnoses of depression, while being pregnant but before receiving their first “Initial Prenatal” visit, as their case/control status was not well-defined.

**Figure 1:**
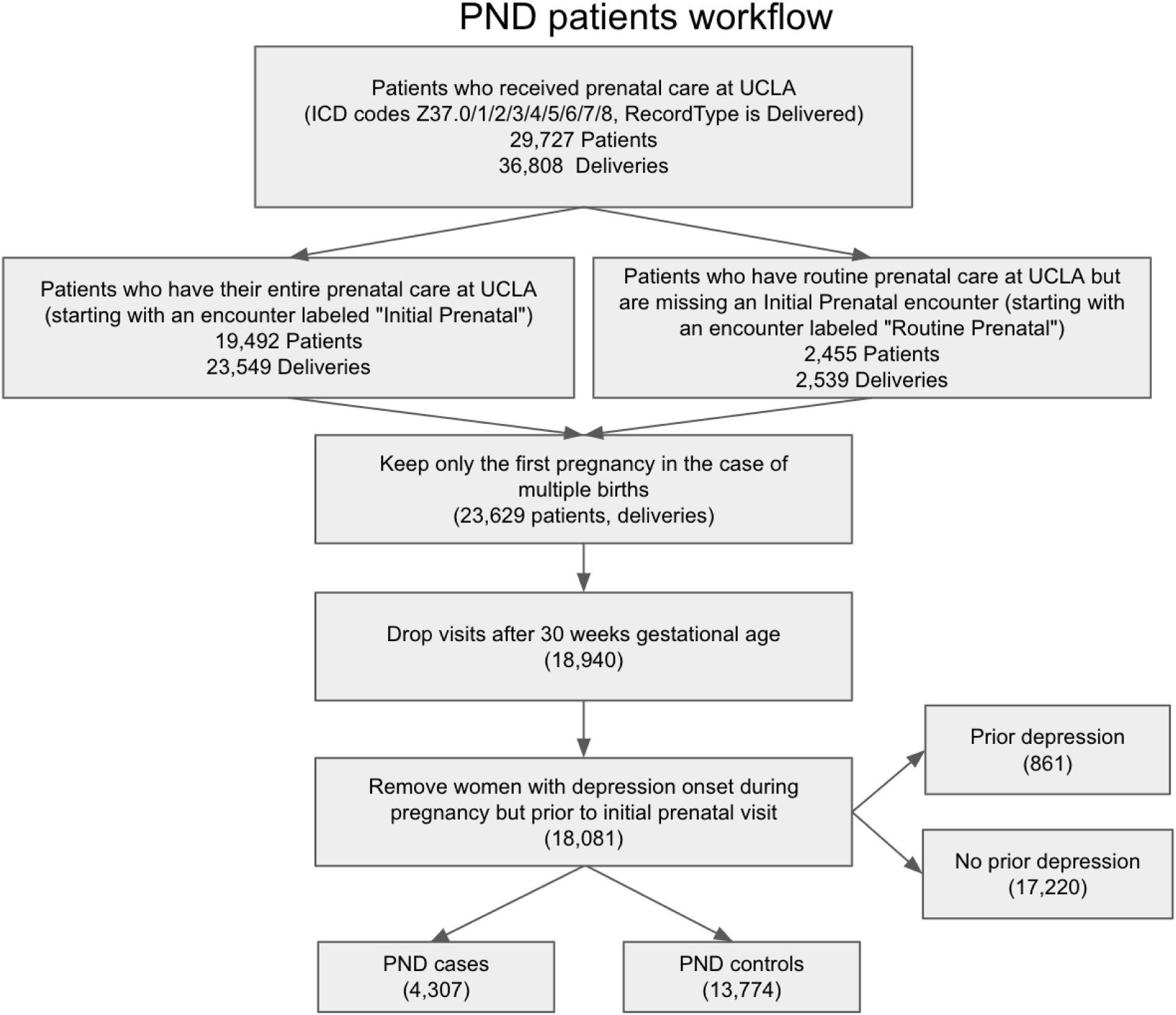
STROBE diagram representing the flow of patients based on inclusion criteria.

### Prevalence and monthly incidence

Case prevalence was defined as the percentage of cases compared to the overall cohort of pregnancies with at least one year of follow up. We further compute incidence during the PND period as the percentage of mothers with an incident PND assignment during each month.

### Predictors

Structured data included demographics, labs values, comorbidities, and medication history as predictors. The following timeframes, classification and imputation strategies were adopted. Demographics were considered at the first prenatal visit. For lab values, the first lab results within 3 weeks of the initial prenatal visit were considered and standardized using z-score normalization (mean = 0, SD = 1) across the cohort. To account for missing lab values, if missingness was less than 50%, we performed mean imputation. For labs with missingness greater or equal than 50%, we included a missingness indicator column. We used the Chronic Conditions Data Warehouse^43^ to define ICD code mapping for comorbidities. The binarized comorbidities from all records before the first visit were considered (presence yes/no). Medications with their corresponding dosage levels that were prescribed before the first prenatal visit were considered and the values were binarized (presence yes/no). The total number of predictors was 194, including 33 demographics, 50 comorbidities, 27 lab values and 84 medication classes. A detailed list of features that were included/excluded can be found in table S3.

### Univariate Regression

To assess the relationship between each variable and PND we performed univariate logistic regression. Each predictor was modeled as an independent variable, with age included as a covariate. Odds ratios, 95% confidence intervals, and p-values were computed for each variable. We adopted a bonferroni correction for multiple testing using a p-value< 0.05/185 based on 185 tests.

### Machine Learning Models for PND Classification

To predict the occurrence of perinatal depression, we employed the following models: Ridge Regression^44^, Gradient Boosting Trees^45^, Random Forests^46^, ExtraTrees^47^. All models were implemented using scikit-learn with default hyperparameters. Ridge Regression used an L2 penalty with default alpha, while the tree-based models (Gradient Boosting, Random Forest, ExtraTrees) used default values for the number of estimators (50), maximum tree depth (None), and minimum samples per split (2).

### Evaluation Metrics

1. **AUROC:** A Receiver Operating Characteristic (ROC) curve was specified for each classification to show the relation between the true positive rate and false positive rate.. We report AUROC values alongside their 95% confidence intervals (CI) to provide a measure of the reliability of these estimates.
2. **AUPRC:** A Precision Recall (PR) curve was specified for each classification to show the relation between precision and recall. The performance of the classifiers was then summarized by the total area under the PR curve (AUPRC), with a higher AUC (between 0 and 1) indicating a better performance of the classification. Similar to AUROC, AUPRC values are reported within 95% confidence intervals to ensure robust evaluation of the classifier’s precision-recall trade-offs.

### Modeling frameworks

We employed a comprehensive modeling framework to assess the predictive performance, robustness, and fairness of our machine learning models in predicting perinatal depression (PND). The framework was structured to account for variations in prior mental health history (1), ethnoracial categories and social determinants of health (2), ensuring a robust and inclusive evaluation.

#### 1. Models With and Without Prior Depression

To understand the impact of prior mental health history on predictive performance, we developed and evaluated models across two distinct cohorts:

a. **Full model (*Cohort_Full*)**: This cohort included individuals with a documented history of depression or mental health diagnoses prior to pregnancy. The inclusion of prior mental health conditions allowed us to leverage these as predictors, yielding models with stronger predictive power but limited generalizability to new-onset cases.
b. **Without Prior Depression (*Cohort_noDEP*)**: This cohort excluded individuals with any prior history of mental health conditions, enabling the development of models tailored to predict PND in populations without pre-existing psychiatric conditions. This approach also highlighted the contributions of biological and socio-environmental factors independent of prior mental health diagnoses.

Finally, we performed a sensitivity analysis with the removal of all individuals with any prior mental health diagnosis **(*Cohort_noMH*).** By comparing the performance of these models, we gained insights into the relative importance of prior mental health history and identified unique predictors for individuals without such a history.

#### 2. Stratification by Ethnoracial Category and Social Determinants of Health

To evaluate the fairness and robustness of our models, we stratified *Cohort_Full* by:

a. Ethnoracial Category: This is an internally defined combination of self-reported race and ethnicity, enhancing clarity in categorization of self-reported race-related data. We included ethnoracial groups with > 1,000 patients each: White, Black or African American, Asian and Hispanic or Latino.
b. Social Determinants of Health: We stratified the data based on three social determinants of health, as follows:

i. SVI Rank: This reflects the social vulnerability of patients. A higher value indicates greater vulnerability, determined by socioeconomic status, household composition, minority status, and housing type. This data aids in identifying areas susceptible to adverse health impacts during emergencies.
ii. BAS Rank: Barriers to Accessing Services: The indicator on Barriers to Accessing Services is meant to capture barriers that increase difficulty in accessing COVID-19 and other general health care services. This indicator is composed of five different variables, namely, non-U.S. citizens, English language barrier, lack of broadband access, lack of health insurance, and vehicles per person, defined as the inverted ratio of vehicles available per person.
iii. Healthy Places Index ^48^: HPI maps data on social conditions that drive health, like education, job opportunities, clean air and water, and other indicators that are positively associated with life expectancy at birth.

For both stratification methods we adopt two approaches: i) We test the model developed on *Cohort_Full* only in a specific group, and ii) We train and test a model in a smaller cohort *Cohort_Group* (e.g., *Cohort_White,* or *Cohort_BASQ1*). This dual approach enables both fairness evaluation as well as the identification of population-specific predictors and performance variations.

### Variable (feature) importance/selection

Variable importance using Random Forests models were calculated using Gini Importance or Mean Decrease in Impurity (MDI)^61^. The MDI relevance of a variable is obtained by calculating how effective the variable is at reducing the uncertainty when creating decision trees. We also used SHapley Additive exPlanations (SHAP) values for local feature importances^49^.

## Results

### PND cohort

Our initial dataset included 36,808 live births from 29,727 mothers. From these, 18,940 had an initial prenatal or routine prenatal visit less than 30 weeks since the start of pregnancy. After excluding 714 mothers who received a depression diagnosis while being pregnant but before receiving their first “Initial Prenatal” visit, and 145 mothers who did not have a complete year of follow up after childbirth, our final cohort included 18,081 patients with 2,601,238 encounters (See Figure 1 detailing our filtering strategy). From these, 13,774 were considered controls and 4,307 cases (23.52% prevalence of PND). Our observed prevalence of PND is high, but falls within the range reported in related literature, spanning from 10-25%^50^. Most index diagnoses occurred close to the delivery date, both before and soon after delivery (Figure 2A). The index diagnosis of PND was non-uniform and included a variety of diagnoses and medication classes. From all PND cases, 60% were included because they received an eligible diagnosis at their index visit, from which 18.7% are included as cases because of Mental disorders and diseases of the nervous system complicating pregnancy, childbirth and the puerperium (O99.3), compared to 39% with Major depressive disorder (F32/F33) and only 12% received a Postpartum depression diagnosis (F53), as their first diagnoses. The remaining 40% of PND cases are included because they start a new antidepressant medication (Figure 2C). From this group, 65% also received an eligible diagnosis within the eligibility window. A sizable subset of PND cases (n=861, 15%) had a reported history of depression prior to childbirth, however, the majority of cases were not previously diagnosed in the UCLA EHR.

**Figure 2.**
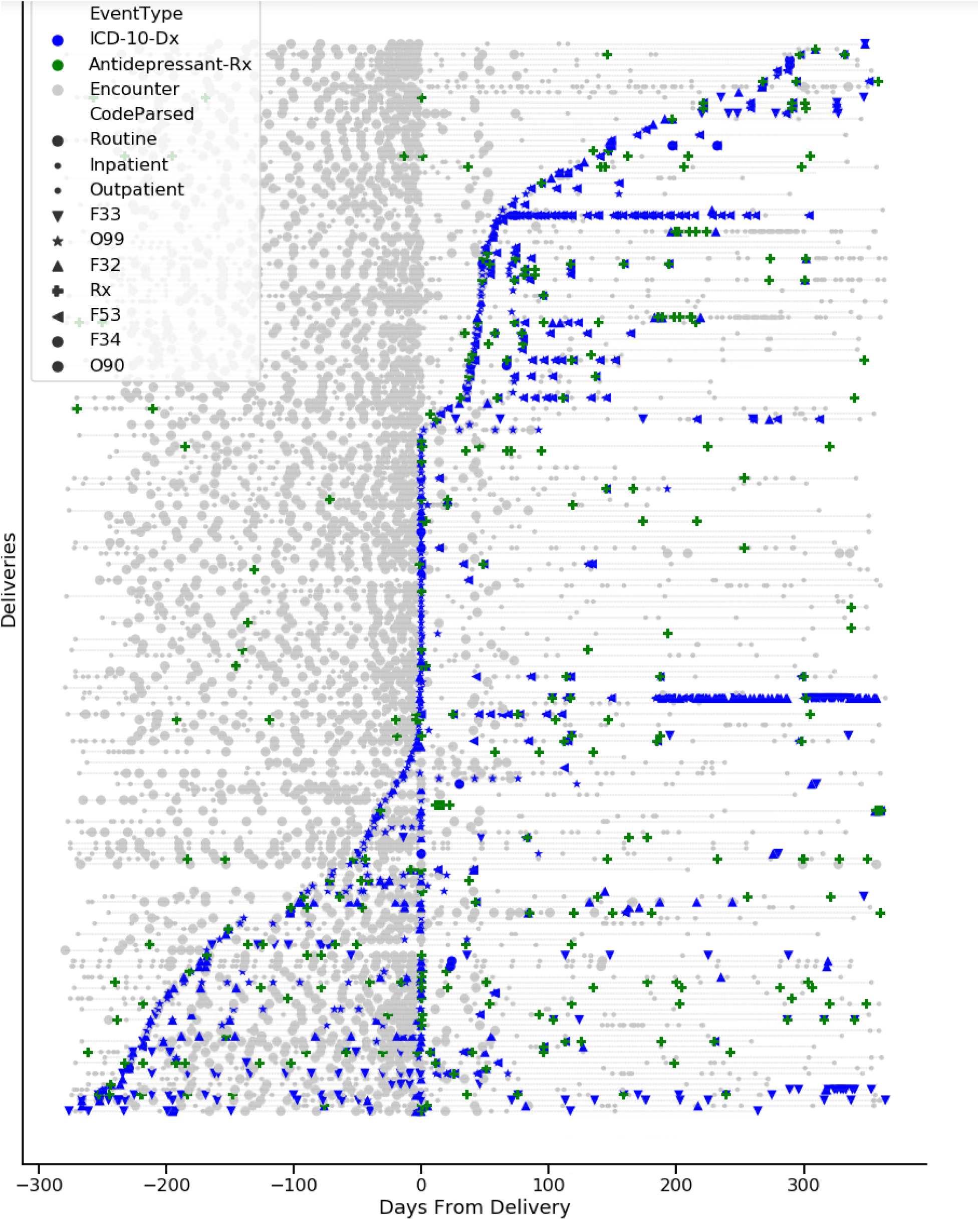
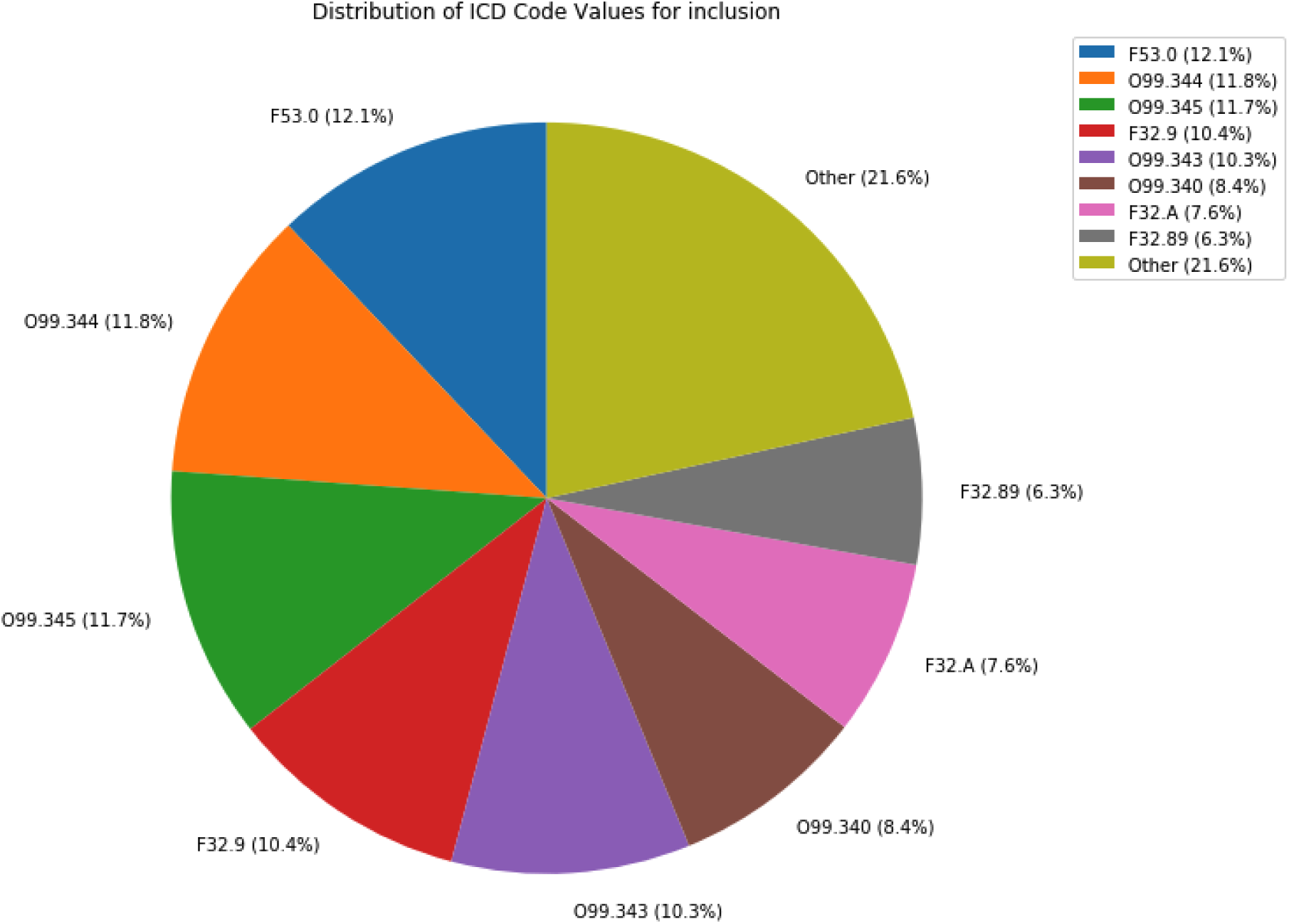
**a:** Healthcare utilization of 200 randomly selected mothers with PND from ATLAS, aligned by delivery date (time 0). Each line indicates one pregnancy and blue colors are PND inclusion by ICD-10, green are PND inclusion criteria by antidepressant prescription. **c:** Inclusion criteria for defining a case of perinatal depression (PND)

We observe the following demographic characteristics of PND cases compared to controls: Compared to controls, PND cases are more likely to identify as White (43% vs 38%), are less likely to be married (78% vs 83%), For the SVI and BAS rank, there is greater incidence of PND in Quartile 4 and the opposite in Quartile 1, differentiating non-marginalized and marginalized populations. In addition to demographics, we observed several significant differences between case and control populations for vitals, labs, medications and comorbidities. First, comorbidities related to prior mental health conditions were significantly different, such as a history of depression (OR=8.7, CI=[7.5,10.2], ADHD (OR=13.8, CI= [10.7,8.3]) and Bipolar Disorder (OR=17.1, CI=[11.1, 26.2]). Several medication classes were recorded at higher frequencies in cases, including those related to mental health, such as antianxiety agents (OR=3.24, CI=[2.88,3.63], antipsychotics (OR=2.4, CI=[2.0,2.7]) and anxiolytics (OR=3.2, CI=[2.9,3.6]). We also observed several blood counts to be different, including red blood cell (RBC) (OR=1.35, CI=[1.31, 1.42]) and white blood cell (WBC) (OR=1.34, CI=[1.30, 1.39]) counts. Finally, vitals such as pulse (OR=1.32 per SD, CI=[1.28, 1.39]) and blood pressure (OR=1.2 per SD, CI=[1.13, 1.27] were different. Aside from many risk factors, the only protective factor we identified was having a partner (OR=0.71, CI=[0.36, 0.65]). Table 1 summarizes various socio-demographic characteristics of this population.

**Table 1:** Characteristics of our cohort for cases and controls.

| Characteristics | PND | Non-PND | P-Value |
| --- | --- | --- | --- |
| **Total** | 4,307 | 13,774 |  |
| **Ethnoracial Category** |  |  | 0 |
| White | 43% | 38% |  |
| Asian | 15% | 22% |  |
| Hispanic or Latino | 20% | 20% |  |
| No Usable Values | 9% | 10% |  |
| Black or African American | 5% | 5% |  |
| Other | 8% | 5% |  |
| **SVI Rank** |  |  | 1.7e-08 |
| Quartile 1 | 23% | 26% |  |
| Quartile 2 | 24% | 25% |  |
| Quartile 3 | 25% | 25% |  |
| Quartile 4 | 28% | 24% |  |
| **BAS Rank** |  |  |  |
| Quartile 1 | 23% | 26% | 0.001 |
| Quartile 2 | 26% | 25% |  |
| Quartile 3 | 25% | 25% |  |
| Quartile 4 | 26% | 24% |  |
| **Healthy Places Index Percentile** |  |  | 0.02 |
| Quartile 1 | 27% | 24% |  |
| Quartile 2 | 24% | 25% |  |
| Quartile 3 | 26% | 25% |  |
| Quartile 4 | 23% | 26% |  |
| **Marital Status** |  |  | 1.17e-15 |
| Married | 78% | 83% |  |
| Single | 16% | 13% |  |
| Significant Other | 3% | 2% |  |
| Other | 3% | 2% |  |
| **Prior depression** | 15.37% | 1.71% | 0 |
| **Prior mental health condition** | 40.4% | 8% | 0 |

**Table 2:** Model performance stratified across different ethnoracial categories according to two methods as follows: 1) the entire dataset was subset on the particular category and trained and tested within that sub-category and 2) the model was trained on the full dataset but tested on held-out individuals belonging to a specific category

| Race | # (total/cases) | Prev % | ROC<br>[95%CI]<sup>1</sup> | PRC<br>[95%CI]<sup>1</sup> | ROC<br>[95% CI]<sup>2</sup> | PRC<br>[95% CI]<sup>2</sup> |
| --- | --- | --- | --- | --- | --- | --- |
| Asian | 3861/628 | 16.43 | 0.74<br>[0.63,0.85] | 0.45<br>[0.37,0.53] | 0.73<br>[0.66,0.8] | 0.45<br>[0.41,0.48] |
| Black or African American | 866/227 | 26.36 | 0.70<br>[0.56,0.81] | 0.47<br>[0.4,0.54] | 0.72<br>[0.64,0.78] | 0.52<br>[0.46,0.53] |
| Hispanic or Latino | 3355/858 | 25.81 | 0.70<br>[0.6,0.81] | 0.48<br>[0.42,0.54] | 0.71<br>[0.64,0.78] | 0.5<br>[0.46,0.53] |
| White | 7249/1882 | 25.96 | 0.73<br>[0.65,0.86] | 0.59<br>[0.52,0.66] | 0.75<br>[0.68,0.83] | 0.56<br>[0.52,0.6] |

### Classification models for the prediction of PND

All model architectures tested had a similar performance range with gradient boosting trees achieving the highest AUROC of 0.75 CI [0.73,0.77] and AUPR of 0.57 CI [0.56,0.57] on the Cohort_Full (Table S6). The Random Forest model provided robust performance with an AUROC of 0.75, CI [0.65,0.84]. However, performance of Cohort_noDEP was reduced AUROC 0.71, CI [0.6,0.8], Figure 1). In a follow-up sensitivity analysis we observed the model performance to further drop on Cohort_noMH (Figure S1, AUROC 0.65, CI [0.53,0.77]). We also performed additional sensitivity analyses including only individuals who received their primary care at UCLA, and found the model performance to be consistent with the full dataset, indicating that we were not biasing our model with some proxy for primary care access (Figure S3).

### Global Feature importances for model interpretability

We computed the global feature importances for the random forest models on Cohort_Full and Cohort_noDEP. The 20 most important variables by gini importance based on the random forest model are shown in Figure 3. For both cohorts, their mental history was the most important factor, followed by social determinants of health (SVI, BAS ranks), followed by patient demographics, vitals and labs. Across both models, the feature importance scores were fairly consistent,with anxiety disorders being the top risk factor for both cohorts.

**Figure 3:**
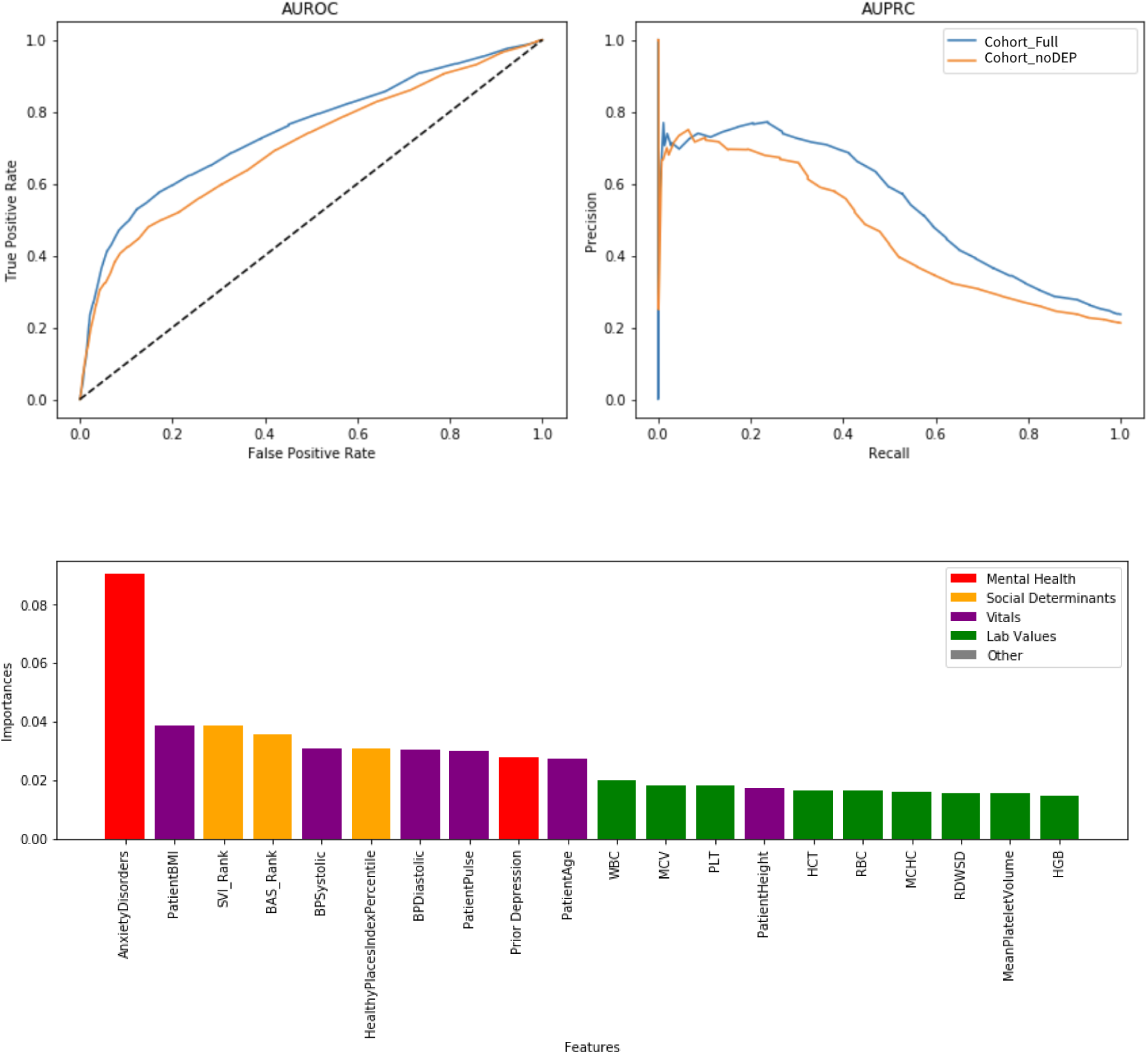
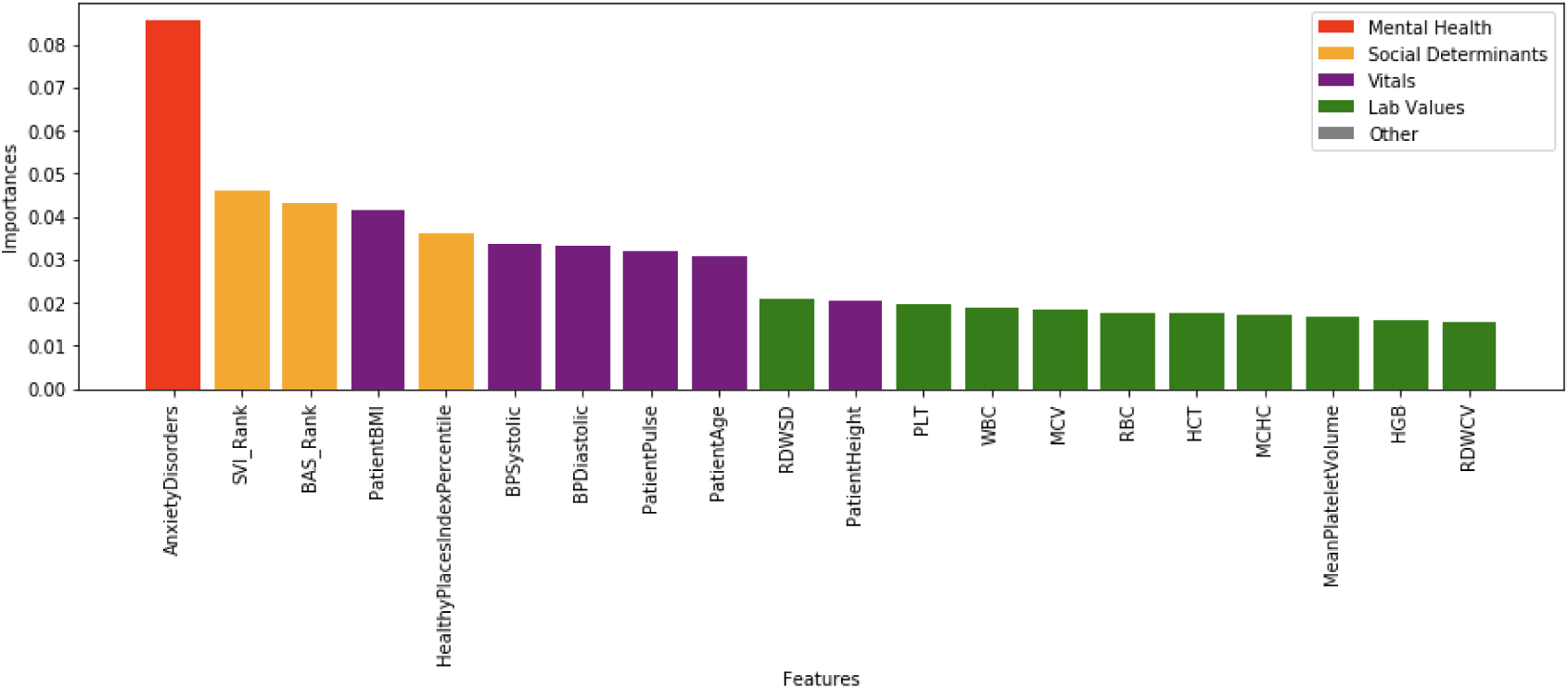
a) Receiver Operating Characteristic (ROC) and Precision-Recall (PR) curves for the model in detecting PND for Cohort_Full and Cohort_noDEP. The model achieved an AUROC of 0.75 and AUPR of 0.55 for the full cohort and AUROC of 0.70 and AUPRC of 0.30 in Cohort_noDEP. b,c) Global feature importances for the models run with and without using prior depression for the top performing random forest model.

### Individual feature importances

To better understand how feature importance varied by individuals, we computed the SHAP values for the random forest model. We found prior mental health diagnosis and treatments: Anxiety Disorders, Prior Depression, Anxiolytics to be the top important features, with a positive value corresponding to a high value on the model output. This was followed by social determinants of health, such as the BAS rank. Healthy Places Index Percentile, and SVI rank. Finally, patient vitals and labs were important, includingBMI, pulse, blood pressure (Diastolic and Systolic), blood counts (RBC, PLT, HCT).

### Model performance stratified by Ethnoracial Category, SVI, BAS Ranks and Healthy Places Index Percentile

To evaluate performance of our model across different Ethnoracial categories, as well as social determinants of health we developed models stratified by these factors. The highest prevalence of PND was observed in the Black or African American group, while the lowest was for the Asian population. Model performance of the general model applied to stratified test data was consistent across the different categories, with overlapping confidence intervals and AUROC scores ranging from 0.70 to 0.75. The highest score corresponded to the White category, while the lowest was for the Black and African American category (Table 3). We computed an equalized odds ratio as another measure of detecting fairness, and found most values to be close to 0.8 (Table S4), which is the standard for fair models^51^, except for Asian vs Black or African American populations (EOR=0.56) (Table S4). For the SVI rank, we found most values to be close to 0.8, with the largest disparity between quartile 1 and 4 (most and least vulnerable populations; table S5). On the other hand, the ratios for BAS ranks were all above 0.8, denoting a fair model with respect to BAS.

### Differential ranking of global feature importances across Ethnoracial Category, and SVI ranks

To better understand the models at a feature level across groups, we next evaluated model feature importance in the stratified models. We computed the top 10 most important features for the category with respect to the largest subset for race (White) and the first quartile for SVI ranks, BAS ranks and Healthy Places Index Percentile. Figure 4 displays the relative ranking of the top features for the remaining categories. The global feature importance remained similar across racial groups, except for prior depression which dropped to rank 29 for black and african americans. In contrast, rankings for social determinants of health remained relatively stable, with none of these features ranking higher than 20 in any quartile when compared to their positions in Quartile 1. It is interesting to note that once we added the other social determinants of health as predictors, the BAS rank and HealthyPlacesIndexPercentile became the top important features. In addition, while anxiety disorders remained the top feature for most races, it was ranked 7 for the Black and African American population, with the SVI rank being the number 1 ranked predictor.

**Figure 4:**
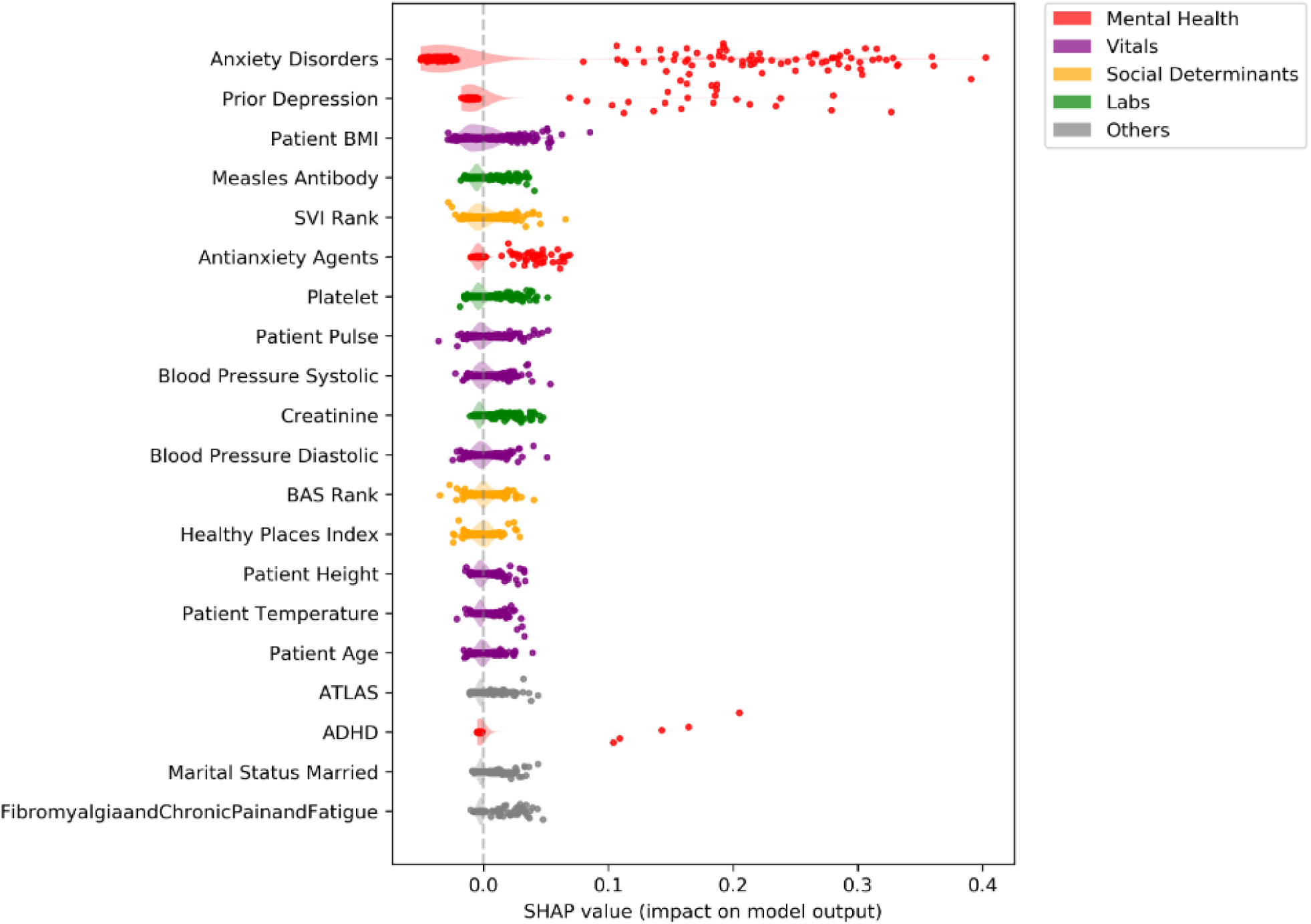
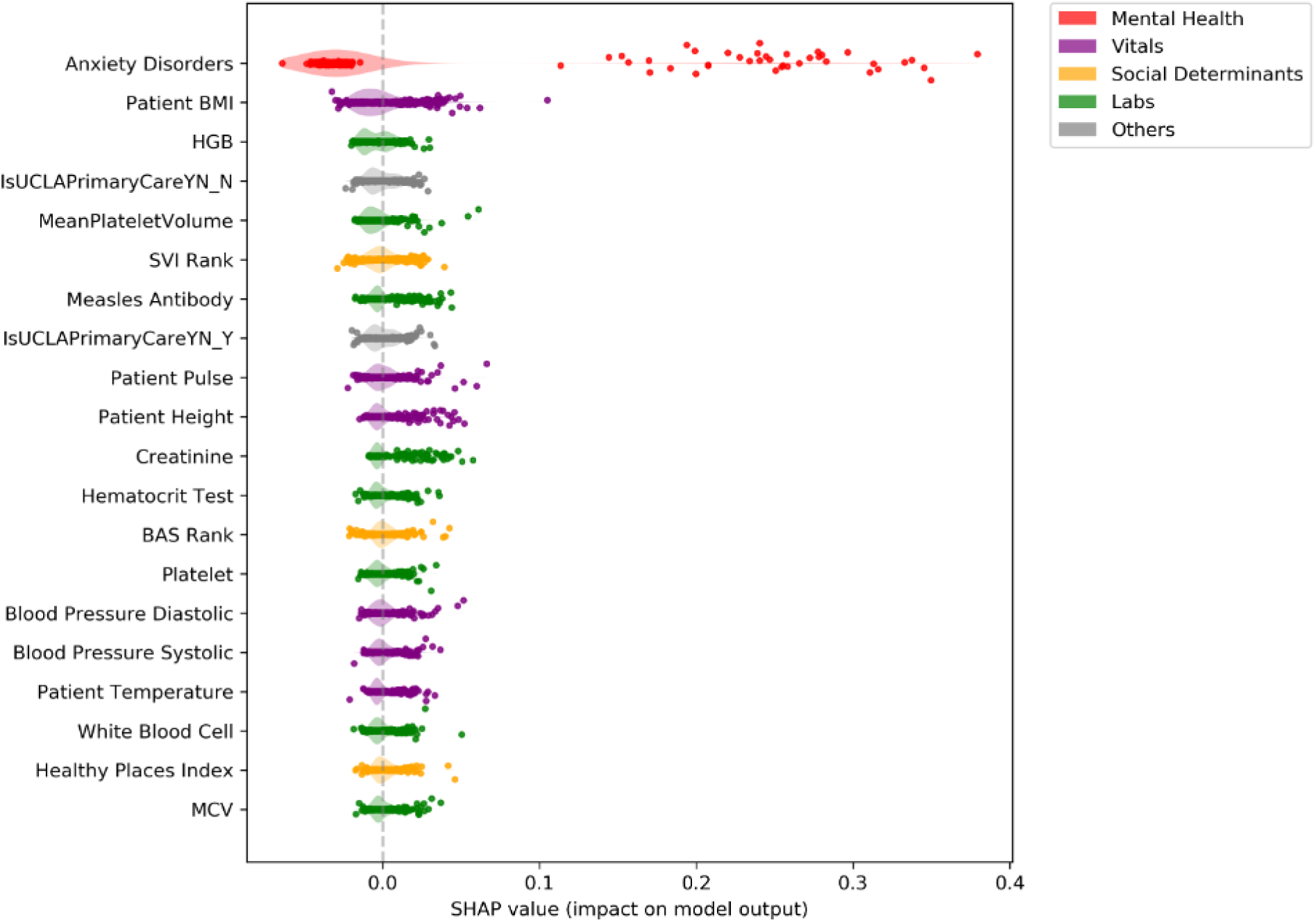
The relative feature importance rankings of the random forest model based on the SHAP value for 500 random test samples for a) with mothers having a history of prior depression and b) without a history of prior depression

**Figure 5:**
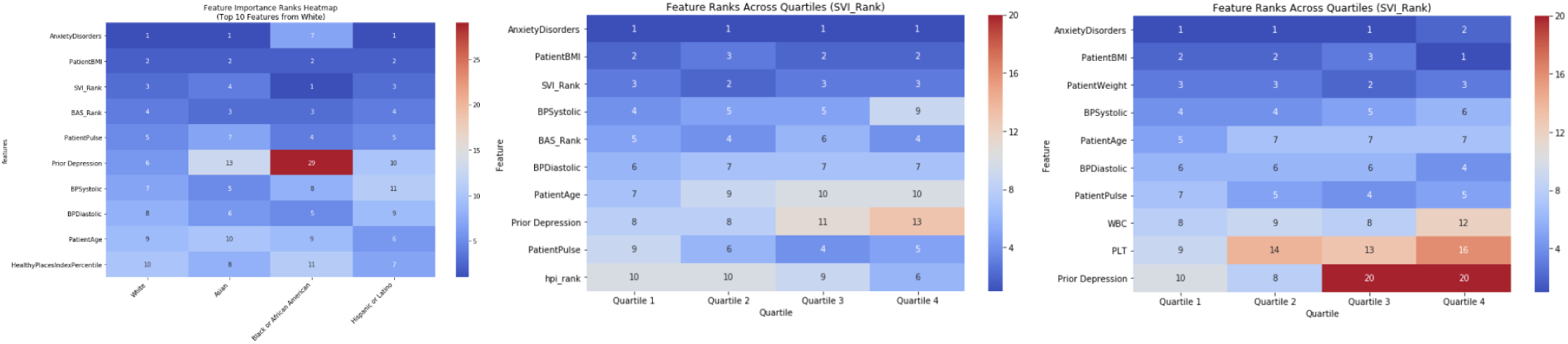
The relative feature importance rankings of the top class with other classes for the a) ethnoracial categories and SVI rank percentile b) prevalence of PND across ethnoracial categories and quartiles c) without and d) with other social determinants of health as features for the random forest model based on the model coefficients. The top class was defined as the class either containing the highest number of samples or the first quartile

## Discussion

We performed early prediction of perinatal depression using electronic health record data from a diverse population of over 18,000 mothers at UCLA. By leveraging a robust dataset and multiple machine learning models, we identify several key predictors and evaluate their performance across diverse populations in a fairness analysis. Our findings not only confirm several previously published risk factors, but also identify several novel risk factors which may hold promise in advancing the early prediction of PND.

### Cohorts stratified by mental health history

Our study is the first to perform prediction on two distinct cohorts based on prior mental health conditions: 1) the full cohort, 2) mothers without a history of prior depression. We also performed a further analysis of mothers without a history of any mental health conditions. Our results show that the predictive performance drops at each level, underscoring the importance of prior mental health conditions as predictors of PND. However, since most PND cases occur in mothers without a prior history of depression, this group of mothers, despite being the most challenging to predict, stands to gain the greatest benefit from an early screening mechanism.

### Novel and known risk factors of PND

In our study, we replicate several known risk factors as features that were both significant in our univariate logistic regression analyses and in the top 20 predicted by the random forest model. These include RBC, WBC counts, age, maternal BMI, blood pressure, and socioeconomic indicators. For Cohort_Full, surprisingly, anxiety was more important than prior history of depression which may signify mild cases of depression that were not captured. Interestingly, the only significant protective factor among demographics was having a partner, suggesting the potential effect of social support in mitigating the risk of perinatal depression.

We also highlight the following novel risk factors:

1. **Lab related markers:** We observed several laboratory test related markers that were important features in the prediction of PND (top 20 predictors), that have not been previously reported to be important features in the prediction of PND. These novel predictors are 1) MPV, 2) RDWSD. These markers could provide new insights into the biological mechanisms underlying PND.
2. **Patient vitals:** We identify patient pulse at first prenatal visit to be among the top predictors of PND. Surprisingly, this feature has never been explored in prior literature. We also found well established risk factors, such as increased age^52^ and increased blood pressure ^53^ to be top predictors of PND. Together, this combination of vitals can improve early prediction of PND.
3. **Social determinants of health**: Our study is the first to use SVI rank, BAS ranks and HPI index to predict the risk of PND. We find these features to be top predictors of PND, signifying the critical role of socioeconomic factors and healthcare accessibility in maternal mental health. We also find a significant increase of English speakers in PND cases, which raises questions about the role of cultural stigma in influencing diagnosis.
4. **Infectious laboratory tests:** We found records of immunity such as the presence of measles, rubella and varicella antibodies to be risk factors. While these associations could be incidental depending on our dataset characteristics, or represent a proxy for access to care, this represents an intriguing avenue for future research.

### Fairness evaluation

We perform an in-depth fairness analysis of our models. While existing fairness studies primarily focus on race and ethnicity, our analysis takes a broader approach. In addition to evaluating fairness across self-reported ethnoracial categories, we also examine three key social determinants of health, namely the SVI rank, BAS Rank, and HPI Percentile, offering a more comprehensive understanding of potential biases in vulnerable populations. Our fairness study highlights several important insights:

**1) Fairness across ethnoracial categories:** In our study, we observed our model performance to vary slightly across ethnoracial groups, with AUROC values ranging from 0.70 to 0.75, and the highest scores observed in the White and the lowest in the Black or African American. Overall, we observed that our models had overlapping ROC confidence intervals and equalized odds scores between ethnoracial groups were close to 0.8, signifying fairness across ethnoracial categories. This is in contrast to related literature where disparities in model fairness are commonly reported^51^. The biggest difference in model fairness through EOR was observed in the case of Asians vs Black or African American population, the two populations with the lowest and highest prevalence of PND, respectively. These findings suggest that our approach is quite robust and equitable in identifying at-risk individuals regardless of their racial or ethnic background. However, we did notice the Asian group to have the lowest TPR and FPR among groups, which could be related to this group having the lowest prevalence of PND.
**2) Diversity in Feature Importance Across Ethnoracial Categories:** While model performance remained stable, the relative importance of certain features varied across ethnoracial groups. For example, anxiety disorders emerged as the top predictor in all ethnoracial categories except for Black or African American. This discrepancy may reflect biases related to social stigma, family support or differences in healthcare access and reporting. These findings emphasize the importance of considering population-specific predictors and addressing potential biases when designing and tailoring interventions for perinatal depression.
**3) Stability of Feature Importance Across SVI rank, BAS rank, and Healthy Places Index Percentile Quartiles:** We observed that the relative feature importance across SVI/BAS/HPI quartiles was mostly stable compared to ethnoracial groups. However, we observed shifts based on the context of other predictors used. For example, for the SVI index quartiles corresponding to the most socially vulnerable individuals, other social determinants of health, such as BAS Rank and Healthy Places Index Percentile, become important predictors, underscoring the importance of the social determinants of health in maternal mental health.

### Limitations

Our study benefits from a large and diverse dataset, thorough methodology, and the integration of machine learning techniques. However, some limitations must be acknowledged. For instance, the absence of the Edinburgh Postnatal Depression Scale (EPDS) in our dataset may have resulted in certain cases of PND being overlooked or inaccurately classified. While our definitions of PND are rigorous, they may not fully encompass the disorder’s complexity and variability. Additionally, this study did not focus on the recurrent nature of PND—whether mothers who experience PND during one pregnancy are more likely to develop it in subsequent pregnancies. More sophisticated analyses strategies, such as imputing the lab based on personal history and gestational age, could be considered. Future work could also explore stratifying depression by the onset time (e.g., onset during pregnancy versus postpartum) and comparing models for different subtypes. We observed anxiety disorders to be the top predictor in most cases, which could be a proxy for undiagnosed prior depression. Exploring these questions in future research could provide valuable insights for clinicians, enabling more proactive and preventive care.

### Conclusions

Our findings emphasize the importance and potential of leveraging EHR data with machine learning models to predict and better understand PND. Early identification of at-risk individuals can pave the way for timely interventions, reducing the morbidity and mortality associated with PND. While our predictions are not yet at the level of clinical utility, we hypothesize that future work including expanded clinical note analyses, genetic risk factors, socio-demographic features, and more complex temporal modeling will improve prediction.

## Data Availability

na

https://docs.google.com/document/d/1oiEq3Cj-IiU42qcMWF2vAjwHC30Kv3c3nsW70aoOUDI/edit?usp=sharing

## Data Availability

Privacy regulations at UCLA prevent our data from being publicly shared.

## Supplementary Figures and Tables

https://docs.google.com/spreadsheets/d/13d3X5QT9NO2_QkBvWmjGQiqkVx4S7oK_x1aRJi8zShU/edit?usp=sharing

**Table S1:** Complete statistics of individuals in our cohort

https://docs.google.com/spreadsheets/d/1WJnHqIB7×3GHRkW2P_Hs0zU9955zFj_-dlz4elSJOHs/edit?us p=sharing

**Table S2:** Medications used to classify PND

**Table S3:** Features dropped in the machine learning analysis.

| Class | Elements | Reference table |
| --- | --- | --- |
| ID's | MomPatientID, MomDeliveryEncounterCSNID, MomEncounterCSNID, BabyEncounterCSNID, PatientEncounterCSNID, ComorbidDepEncounterID | All |
| Other Identifiers | EncounterType, IsEDVisit, IsInpatient, LaborStatus, Department, DischargeDisposition, DepartmentAbbreviation, DiagnosisEventType | label_df |
| Dates | DaysToOrder, ICDToDeliveryDays, IsICU, NonICUDays, ICUDays, LengthOfStay, InitialEncounterToDeliveryDays, LastVisitAfterInitialDays, DeliveryContactDate, InitialVisitDate, EarliestICDRecordDate, LastVisit, EncounterContactDate, 'EarliestAntidepressantsRecordDate', RxToDeliveryDays, StartDate | label_df |
| Demographics | White, Black, Latinx, IsUCLAPrimaryCareYN, PreferredLanguage, Ethnicity, FirstRace, ThirdRace, BenefitPlanCategory, API, EnglishPreferred, Other, SecondRace, SpanishPreferred, bank_sample_id, condition | demographics |
| Redundant | DepressionAndDepressiveDisorders, Sex, Female, BirthDate, Antidepressants, DeathDate, 'NoReligion' | comorbidities |
| Use to stratify, drop at the end | EthnoracialCategory, SVI_Rank, BAS_score, PND | demographics |
| Same value (>99.9% of the time) | Z37.1, Z37.4, Z37.5, Z37.6, Z37.7<br>AndrogenAnabolic, BiologicalsMisc, DigestiveAids, MiscRespiratory, Sulfonamides, NGONPCR, AlzheimersDementia, AutismSpectrumDisorders, CerebralPalsy, ColorectalCancer, EndometrialCancer, LungCancer, MuscularDystrophy, OtherDevelopmentalDelays, ProstateCancer<br>SensoryBlindnessandVisualImpairment | comorbidities |

**Table S4:** Equalized odds ratio for ethnoracial categories a)

| Comparison | TPR Ratio | FPR Ratio | Equalized Odds Ratio | Fair? ($\geq 0.8$) |
| --- | --- | --- | --- | --- |
| Asian vs Black/African American | 0.63 | 0.56 | 0.56 | No |
| Asian vs Hispanic/Latino | 0.72 | 0.63 | 0.63 | No |
| Asian vs White | 0.68 | 0.7 | 0.68 | No |
| Black/African American vs Hispanic/Latino | 0.88 | 0.89 | 0.88 | Yes |
| Black/African American vs White | 0.91 | 0.8 | 0.8 | Yes |
| Hispanic/Latino vs White | 0.95 | 0.89 | 0.89 | Yes |

**Table S5:** Equalized odds ratios for a) SVI and b) BAS ranks.

| **SVI Comparison** | **TPR Ratio** | **FPR Ratio** | **Equalized Odds Ratio** | **Fair? ($\geq 0.8$)** |
| --- | --- | --- | --- | --- |
| Quartile 0 vs Quartile 1 | 0.93 | 0.69 | 0.69 | No |
| Quartile 0 vs Quartile 2 | 0.93 | 0.69 | 0.69 | No |
| Quartile 0 vs Quartile 3 | 0.89 | 0.57 | 0.57 | No |
| Quartile 1 vs Quartile 2 | 0.99 | 1 | 0.99 | Yes |
| Quartile 1 vs Quartile 3 | 0.83 | 0.84 | 0.83 | Yes |
| Quartile 2 vs Quartile 3 | 0.84 | 0.83 | 0.83 | Yes |

| **BAS Comparison** | **TPR Ratio** | **FPR Ratio** | **Equalized Odds Ratio** | **Fair? ($\geq 0.8$)** |
| --- | --- | --- | --- | --- |
| Quartile 0 vs Quartile 1 | 0.97 | 0.94 | 0.94 | Yes |
| Quartile 0 vs Quartile 2 | 0.93 | 0.94 | 0.93 | Yes |
| Quartile 0 vs Quartile 3 | 0.96 | 0.84 | 0.84 | Yes |
| Quartile 1 vs Quartile 2 | 0.9 | 1 | 0.9 | Yes |
| Quartile 1 vs Quartile 3 | 0.93 | 0.8 | 0.8 | Yes |
| Quartile 2 vs Quartile 3 | 0.97 | 0.8 | 0.8 | Yes |

**Table S6:** Predictive performances (ROC, PRC) using various machine learning models.

| Model | AUROC | AUPRC |
| --- | --- | --- |
| Random Forest | 0.75 | 0.55 |
| Gradient Boosting Trees | 0.75 | 0.57 |
| ExtraTrees | 0.74 | 0.54 |
| Ridge Regression | 0.75 | 0.56 |

**Figure S1:**
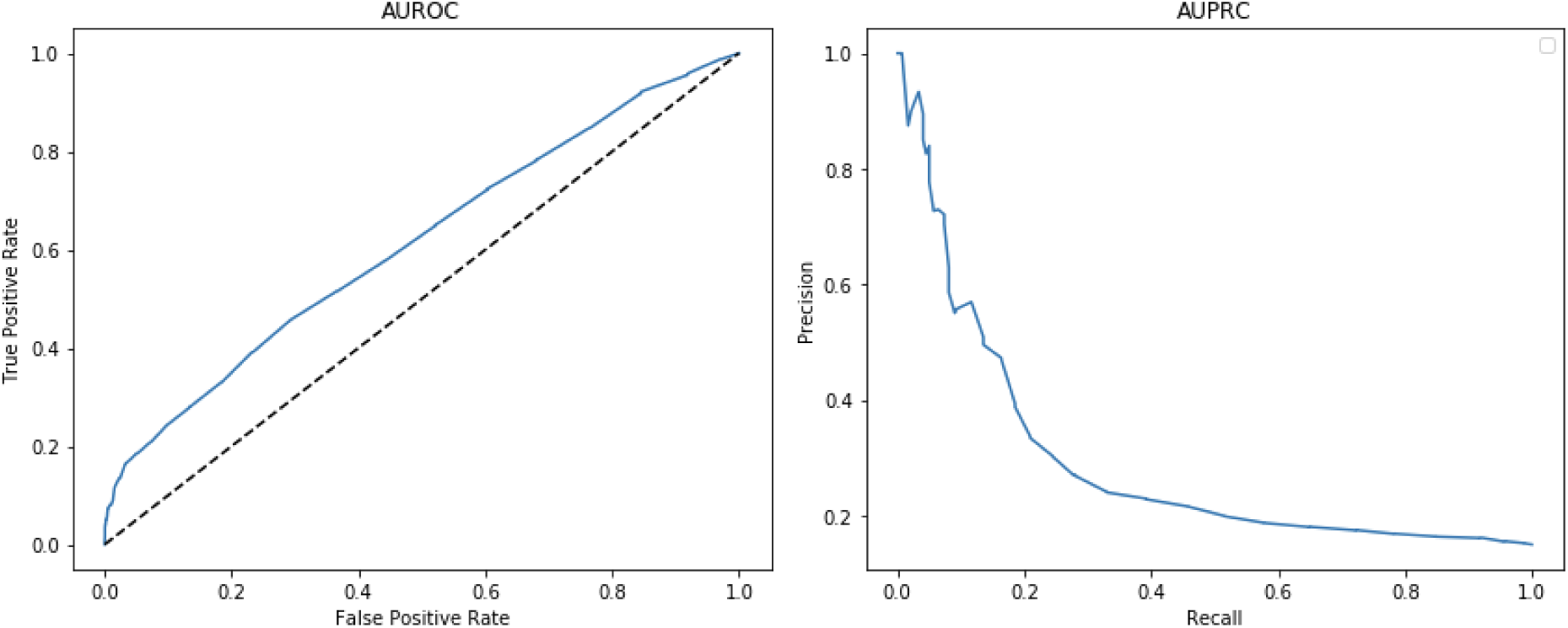
Model performance for predicting perinatal depression with a population with no prior history of any mental illness. All individuals to have received a F code in the past were removed from this analysis

**Figure S2:**
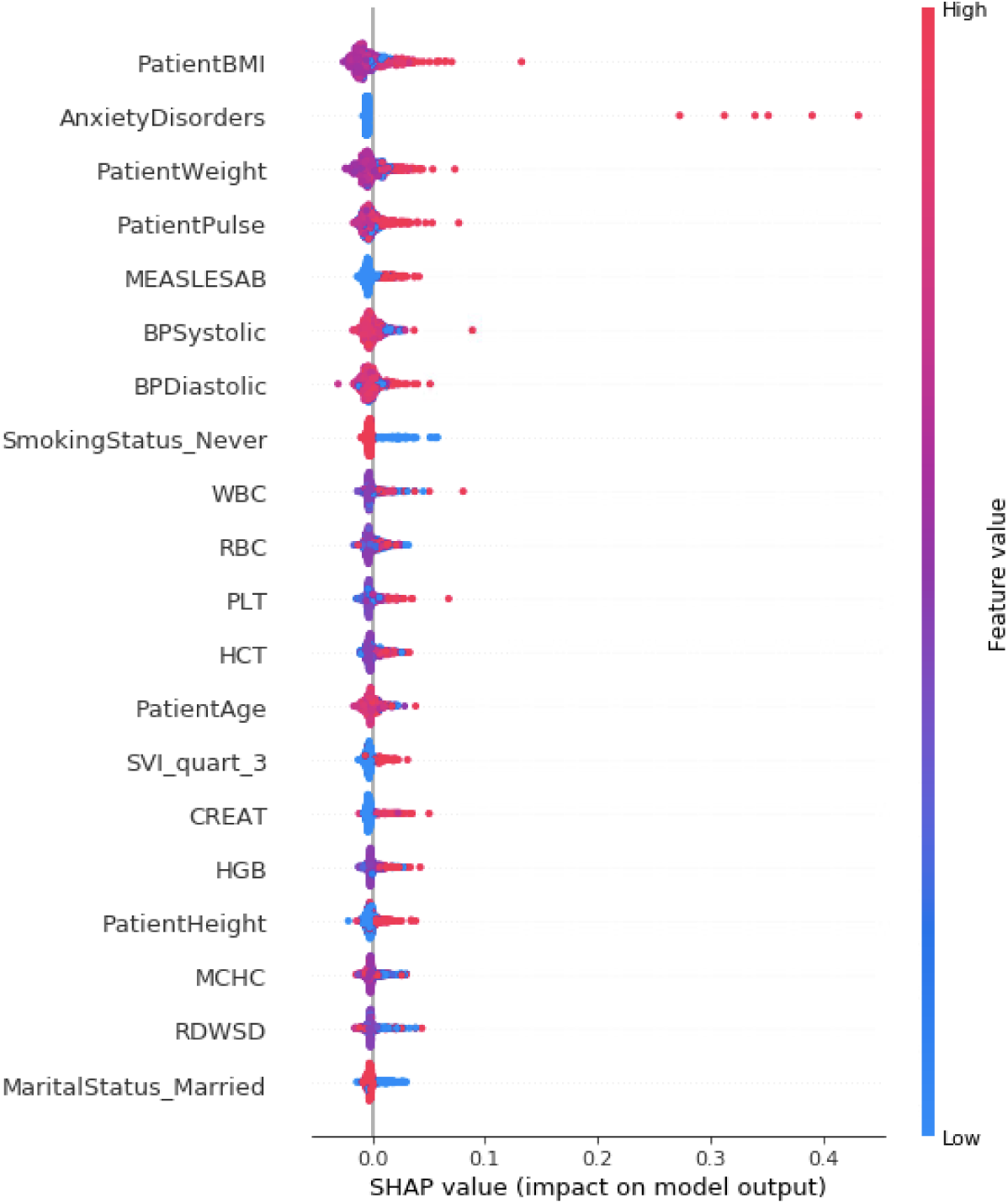
SHAP values from predicting perinatal depression with a population with no prior history of any mental illness. All individuals to have received a F code in the past were removed from this analysis.

**Figure S3:**
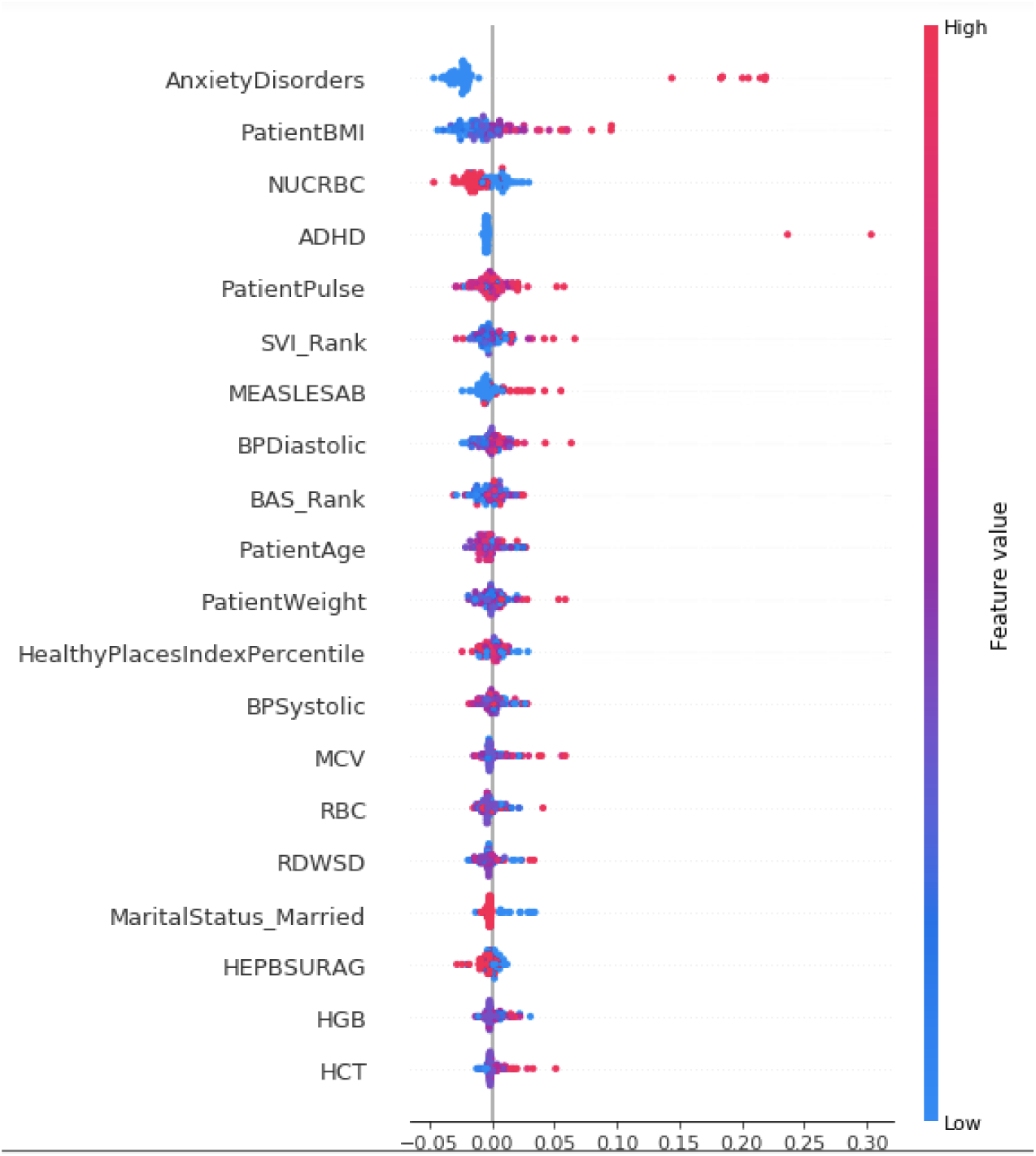
SHAP values from predicting perinatal depression with a population with primary care at ucla. Individuals with prior depression were not included.

**Figure S4:**
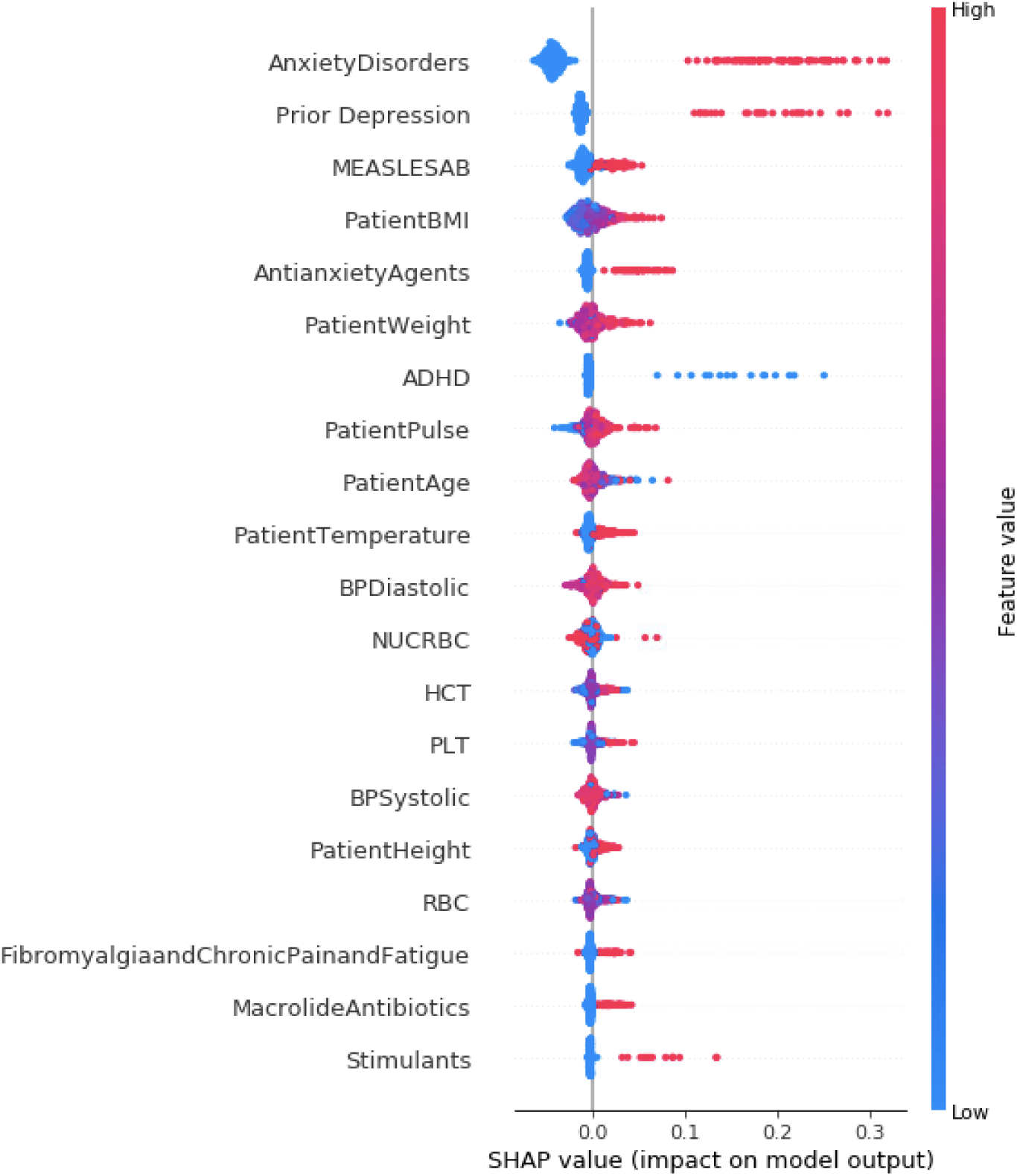
SHAP values from predicting perinatal depression with a population with primary care at ucla. Individuals with prior depression were included.

**Figure S5:**
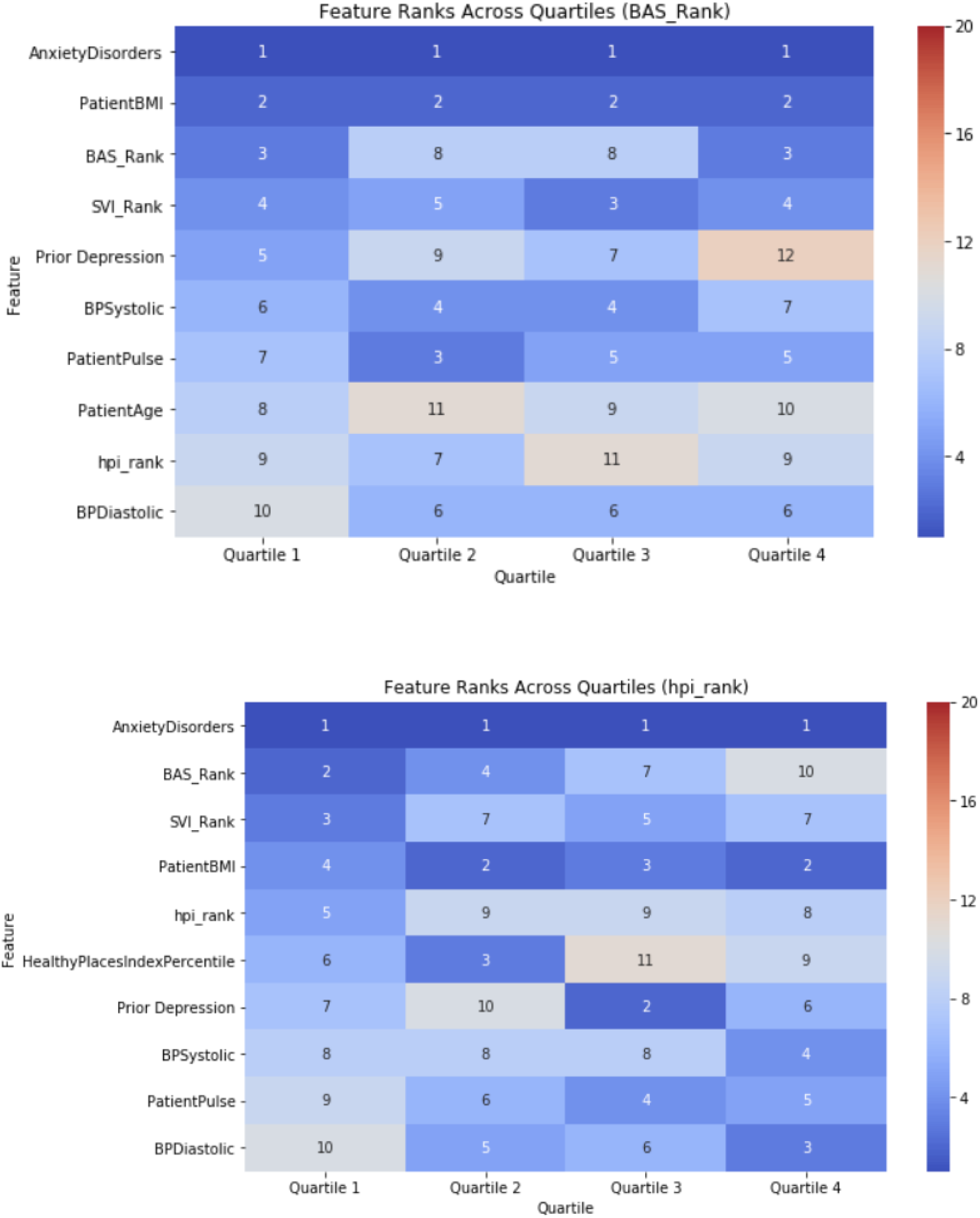
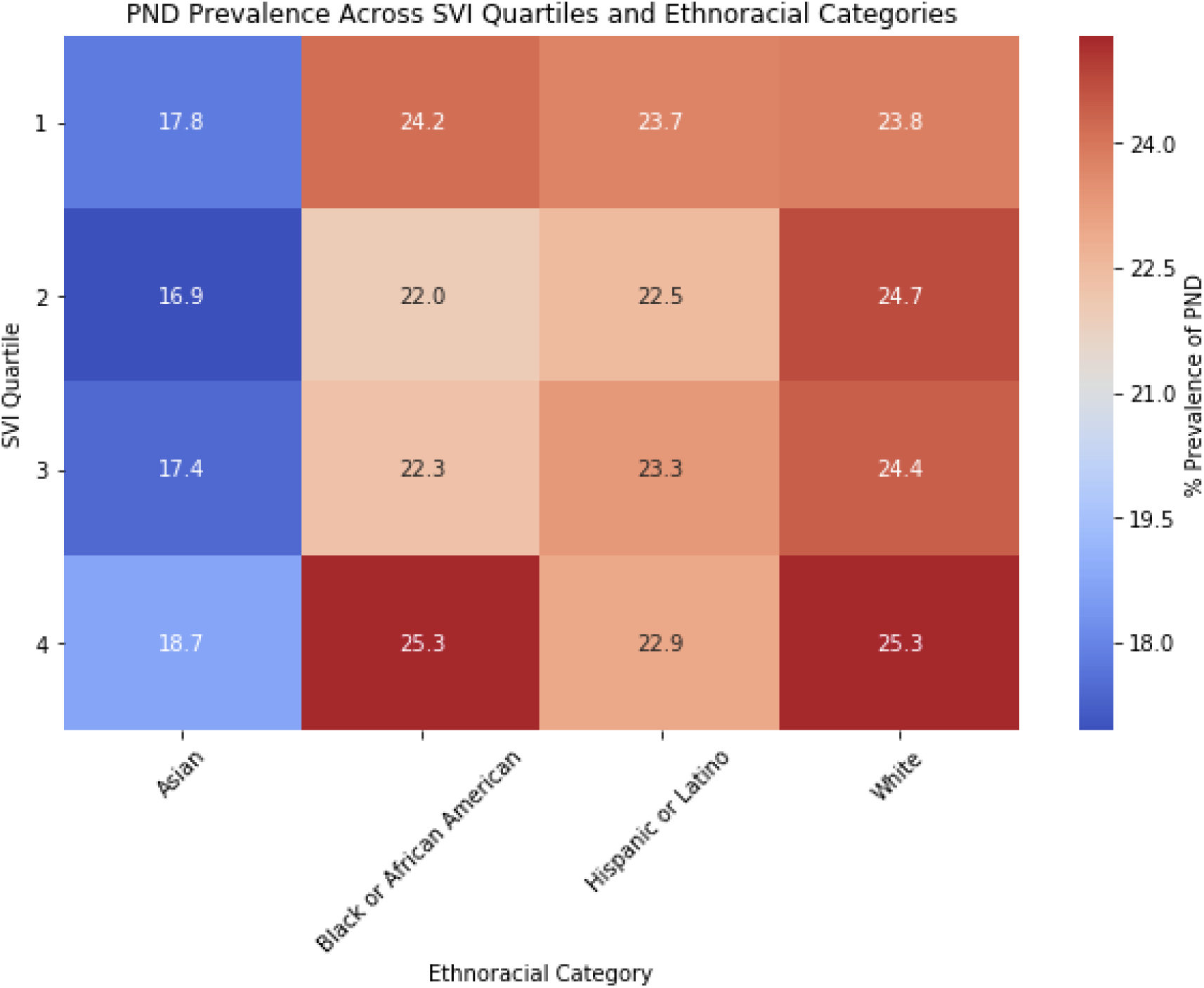
The relative feature importance rankings of the RANDOM FOREST model based on the a) BAS Rank Quartiles b) Healthy Places Index Percentile quartiles and c) SVI quartiles vs ethnoracial categories.

